# Online randomised trials with children: A scoping review protocol

**DOI:** 10.1101/2022.04.15.22273742

**Authors:** Simone Lepage, Aislinn Conway, Noah Goodson, Paul Wicks, Declan Devane

**Affiliations:** HRB-Trials Methodology Research Network, School of Nursing and Midwifery, National University of Ireland, Galway, Galway, Ireland; Better Outcomes & Registry Network (BORN) Ontario, Children’s Hospital of Eastern Ontario - Ottawa Children’s Treatment Centre, Ottawa, Ontario, Canada; Children’s Hospital of Eastern Ontario Research Institute, Ottawa, Ontario, Canada; Faculty of Medicine, University of Ottawa, Ottawa, Ontario, Canada; Data & Analytics, Thread Research, Tustin, California, USA; Wicks Digital Health, Lichfield, Staffordshire, United Kingdom; Evidence Synthesis Ireland; Cochrane Ireland

## Abstract

**Introduction:** This scoping review will determine how online, randomised trials with children are conducted. The objectives of the review are: (a) to determine what methods and tools have been used to create and conduct online trials with children and (b) to identify the gaps in the knowledge in this field.

Over the last decade, randomised trials employing online methods have gained momentum. Decentralised methods lend themselves to certain types of trials and can offer advantages over traditional trial methods, potentially increasing participant reach and diversity and decreasing research waste. However, decentralised trials that have all aspects of the trial exclusively online are not yet common, and those involving children even less so. This scoping review will describe and evaluate the methods used in these trials to understand how they may be effectively employed.

**Methods:** Methods are informed by guidance from the Joanna Briggs Institute and the Preferred Reporting Items for Systematic Reviews and Meta-analyses extension for scoping reviews. The search strategy was developed in consultation with an information specialist for the following databases: MEDLINE, CENTRAL, CINAHL, and Embase. Grey literature searches will be completed with the consultation of experts in decentralised trials and digital health using internet searches and suitable trial registries. Once identified, included full-text studies’ references will be manually searched for any trials that may have been missed. We will include randomised and quasi-randomised trials conducted exclusively online with participants under the age of 18 published in English. We will not limit by country of conduct or date of publication. Data will be collected using a data charting tool and presented in text, graphical, and tabular formats.

**Ethics and Dissemination:** Ethical approval is not needed since all data sources used are publicly available. The review will be available as a preprint before publication in an open-access, peer-reviewed journal.

## Introduction

Digital technologies are increasingly important in most aspects of daily life. Health research is especially well poised to benefit from the possibilities these technologies offer. In the last decade, there has been rapid growth in randomised controlled trials (RCTs) and clinical trials using remote and digital methodologies [1-3]. The COVID-19 pandemic has spotlighted these remote methods by emphasizing how useful and innovative they can be. While traditional or centralised trials remain invaluable and necessary, decentralised trials may offer increased or different benefits to trial participants, researchers and health professionals. Decentralised trials include any study where data are collected in an alternative manner to a centralised trial. They often use alternative data collection locations, such as mobile or local health clinics, employ digital technologies, for example, wearable devices, or leverage online methods. Decentralised trials can potentially reach people in geographically remote areas, recruit larger and more diverse populations, improve trial participant retention, and may decrease trial costs and research waste [2, 4, 5].

Recruitment and retention of trial participants are two of the most common hurdles faced by research teams conducting trials with adults [6]. Paediatric trials often encounter the same hurdles as those involving adults and have additional challenges, especially those of a logistical and ethical nature. In their Cochrane systematic review discussing strategies to improve recruitment in trials, Treweek et al identified the lack of, and critical need for, the “development and evaluation” [6(p22)] of strategies for recruitment to paediatric trials. Pica and Bourgeois [7] found that the two most common reasons researchers conducting paediatric RCTs cited for discontinuation or non-publication of a trial were participant recruitment and “conduct problems” [7(p3)], which they defined as technical or logistical issues in completing the trial. They found that 19% of registered paediatric trials went unfinished, and a further 30% went unpublished, resulting in substantial research waste and undue burden to their participants [7]. RCTs can be onerous for participants, which can affect their willingness to enrol or remain in them [8]. Consent forms for RCTs are customarily long documents written in legal jargon, and participants often do not fully comprehend or read them [9]. Given the opportunities online trials offer both participants and researchers for increased engagement and recruitment, and decreased research waste, it is important to examine how these tools can and are directed at paediatric research.

Price *et al* [10] published a systematic review of online trials with adults in 2019, outlining the need for more rigorous reporting of methodologies, but after a preliminary search of MEDLINE, Epistemonikos, and the Cochrane Database of Systematic Reviews, no such reviews, systematic or otherwise, were identified for online trials involving children. Overall, paediatric trials are published less often than adult trials despite paediatric disease making up 26.2% of all-cause disability-adjusted life years [11]. Groff *et al* [11] found of 4146 published RCTs, only 14.2% (591) were conducted with children and 18.3% (761) enrolled both children and adults. This may be because working with children introduces the added challenges of gaining consent from caregivers and children, recruiting children from non-traditional family groups where all caregivers must give consent, and differing international laws regarding the age of health or digital consent. As exclusively online, decentralised trials are still not common, and those with children even less so, there is a gap in the literature describing the methods and processes employed by them. This scoping review will identify, describe, and characterize how online, decentralised trials with children are conducted. The objectives of this scoping review are: (a) to determine what methods and tools have been used to create and conduct online trials with children and (b) to identify the gaps in the knowledge in this field.

Early in the field, Bull *et al* [12] conducted a randomised trial via social media with 16-to 25-year-olds. The researchers used multiple forms of recruitment, including online personal channels, blogs, websites, and in-person recruitment efforts and then conducted the rest of the trial online [12]. This type of research is termed ‘hybrid’, meaning it is conducted partially online and partially through other channels. Systematic reviews have reported on internet-based trials developed for young people and adolescents to address tobacco-, alcohol-, and drug-use, but many of these have an offline component, such as recruitment or consent acquisition [4, 13]. Khan *et al* [14] published a systematic review of web-based interventions for children and young people with neurodevelopmental disorders and identified ten papers that met their inclusion criteria. Most of these trials were also hybrid as they recruited predominantly from clinics, but they demonstrate the trend towards decentralised trial development [14].

While there are numerous publications from ‘hybrid’ trials, a growing body of work is being done entirely online. Notably, Rhodes *et al* [3] published recently on three of the studies they are developing or have already completed via their website ‘Discoveries Online’ [15]. The paper describes their methods for overcoming the barriers involved in conducting their online studies using a webcam and participants’ guardians uploading their data for analysis by the research team [3]. Similarly, both MIT and Yale have developed websites for parents and their young children to take part in online trials, which can be accessed at the ‘LookIt’ [16] and the ‘Child Lab’ websites [17], respectively. The website ‘Children Helping Science’ [18] hosts online trials from multiple groups, mostly academic, for parents and children to access and take part in. Gijbels *et al* [19] also reported on three ‘researcher-moderated’ virtual studies discussing the various elements needed to produce valid, trustworthy results. Although some trials will never lend themselves to being entirely online, the number that are accomplishing this is swiftly increasing.

## Methods

Given the exploratory nature of our objectives, a scoping review is the most appropriate approach for this study [20]. The protocol’s organization and development are informed by the Preferred Items for Systematic Reviews and Meta-Analyses extension for Scoping Reviews (PRISMA-ScR) [21], the JBI Manual for Evidence Synthesis’ scoping review chapter [22], and Peters *et al* [23] and addresses the following steps: identifying a research question, identifying relevant studies, selecting studies, charting the data, and collating, summarising and reporting results. The preliminary PRISMA-ScR checklist is included here (S1 Checklist). This scoping review protocol is registered with Open Science Framework (https://osf.io/ha3kf).

### Identifying a Research Question

This scoping review intends to answer the question, “How have online, randomised trials been conducted with children?” The question was structured using the PCC mnemonic (Population, Concept, Context). The population is children, the concept is randomised and quasi-randomised trials, and the context is internet-based interventions. There had been no synthesis of these studies published at the time of our initial scoping search. For this study, children are defined as anyone under 18 years of age as per the UN General Assembly on The Rights of The Child [24]. We will describe the processes of recruitment and retention, ethics, randomisation, intervention, and data collection and analysis within included trials. Once described, we will categorise and summarise these methods.

### Identifying Relevant Studies

The search strategy was constructed following the guidance of the JBI Manual for Evidence Synthesis [22] and the PRISMA extension for Scoping Reviews [25]. One author (SL) undertook an initial scoping search of MEDLINE and the Cochrane Central Register of Controlled Trials (CENTRAL) to identify articles on the topic. The keywords, free text and indexed terms of relevant articles were used to develop a preliminary search strategy for MEDLINE (Ovid). This draft was then edited and refined by a different author (AC), who is trained as an information and knowledge translation specialist. The goal of the search strategy is to balance both specificity and sensitivity to capture as many relevant studies as possible and work within the constraints of time and resources available for this review.

The databases used for our search are MEDLINE (Ovid), Embase (Elsevier), CINAHL (Ebsco), and the Cochrane Central Register of Controlled Trials (CENTRAL). This is a new and rapidly developing field; therefore, we will perform a grey literature search of the following trial registries: the World Health Organization’s (WHO) International Clinical Trials Registry Platform (ICTRP), the EU Clinical Trials Register, and the NIH Clinical Trials Register. Two authors (NG & PW), experts in the fields of decentralised trials and digital health, respectively, will consult on the grey literature searches to identify trials not captured by traditional databases. We will also search Google and Google Scholar and manually screen the reference lists of all included trials for additional studies. To identify publications still in the review process we will search the preprint servers: medRxiv, JMIR Preprints, HRB Open Research, and Advance from SAGE.

The current search strategy, including all identified keywords, free text, and thesauri terms (MeSH, Emtree, CINAHL headings), has been translated for Embase, CINAHL and CENTRAL and will be adapted for all other information sources iteratively. We initially used the Polyglot translator tool to assist with translations of the search strategy from one database to another [26]. This automated translator has shown that it can decrease the time spent translating search strategies from one database to another [27]. We amended the translation in a two-step process where one author (SL) made the first modifications and then a second author (AC) further revised the translations. The cost of translation services is not within the budget for this review, so included studies will be limited to those published in the English language. We will not limit included studies by date or country of conduct. The search strategy devised for MEDLINE is included here (S2 Appendix) but may be modified as the review develops. All modifications to this strategy will be reported in the final review.

### Selecting Studies

Following the search, we will import all identified citations into EndNote20 [28] reference management software, collate the libraries, and remove duplicates. The titles and abstracts will then be uploaded into the SysRev [29] digital tool for screening. Two authors will perform an initial pilot test for inter-rater reliability (IRR) on 100 titles and abstracts. Inter-rater reliability will be calculated using Cohen’s kappa [30]. Once an agreement of 0.75 is achieved, the remaining titles and abstracts will be screened independently by one author (SL). The decision for a single reviewer to screen the remaining titles and abstracts was made in the interest of resource conservation. Taking into consideration an IRR of 0.75, the single reviewer’s familiarity with the objectives of the review, and the inherent exploratory nature of scoping reviews, we believe this is a valid and robust approach.

The next step in the selection process will be to identify full-text studies that meet the inclusion criteria. SysRev [29] will also be used for full-text screening. A pilot test for this step will be completed independently by two authors for inter-rater reliability using Cohen’s kappa [30]. We will screen 50 full-texts and inclusion criteria may be modified and discussed during this process, and a third author consulted if needed. Once an IRR of 0.75 or greater is reached, one author (SL) will screen the remaining full-text studies independently in the interest of resource and budgetary constraints. Inter-rater reliability tests will be completed for 50 full text studies for every 1000 studies screened.

#### Inclusion Criteria

♦ Randomised and quasi-randomised trials
♦ Trials with participants under the age of 18
♦ Trials enrolling participants under the age of 18 at trial commencement who turned 18 during the trial
♦ Trials where participants were an age-range cohort (for example 16–25-year-olds) and ≥60% of participants were under the age of 18
♦ Trials that enrolled caregiver/child(ren) dyads or groups where the children were active participants who contributed data
♦ Entirely online trials using digital technologies connected to the internet in each phase of the trial (For example, recruitment occurred via social media site advertisements and notice-board advertisements in primary care physicians’ offices which directed participants to a study website to enrol)
♦ Trials that implemented self-managed interventions that were offline, where the participants uploaded data via the internet (For example, if a video of a child
♦ performing a task is uploaded and then assessed by the researchers, or if a child performs 10 jumping jacks before doing their homework and then reports online on their ability to focus on their homework)
♦ Trials that advertised digitally to a school, clinic, or similar, inviting children and/or caregivers to participate on an individual basis

#### Exclusion Criteria

♦ Trials where both caregivers and children were participants, but data was only gathered from caregivers (For example, where a behavioural intervention is implemented and the caregiver submits observations online but the child has no data input)
♦ Trials that employed any intervention that required clinical or researcher measurements offline or in-person
♦ Studies that used online interventions in educational, clinical, or similar group settings where the participants were recruited or interventions were overseen in-person (For example, an entire class takes part in a mandatory online intervention during school hours with the oversight of an educator or researcher)
♦ Posters and conference abstracts due to the paucity of method reporting

Exclusions of full-text trials and their justifications will be recorded and reported. The search results will be reported in full in the scoping review and presented in a Preferred Reporting Items for Systematic Reviews and Meta-analyses extension for scoping reviews (PRISMA-ScR) flow diagram [25].

### Data Charting

We will chart the data from included publications with a data charting tool developed by the authors (Table 1). Two reviewers will pilot and modify the tool using five randomly selected studies, iteratively. The reviewers will then discuss and agree on modifications needed to develop a final data charting tool. In the interest of resource conservation and given review purpose, a single reviewer will chart the remaining studies’ data.

**Table 1.** Preliminary Data Charting Tool.

| Domain | Description | Domain | Description |
| --- | --- | --- | --- |
| Author | First author only | Cross-platform compatibility | Was there cross-platform compatibility? If so, describe. |
| Title | Full title of publication | Tools used for data protection processes | What methods/tools were used for data protection? |
| Year of Publication | Year of publication | Tools used for consent and assent | What methods/tools were used for consent/assent acquisition? |
| Country(ies) of Conduct | If stated, where did the authors conduct their study? | Consent/assent validation | Was there an informed consent/assent validation performed? If so, describe. |
| Country(ies) of Participants | If stated, what country(ies) were the participants living in? | Interventions | Describe the intervention: (type of participant) received (type of treatment) in (type of setting). |
| Type of study | RCT, Quasi-randomised trial | Episodes/frequency of engagement | How many interactions did participants have with the platforms? How frequent were the interactions? |
| Type of publication | Peer-reviewed journal, preprint, other. If other, please describe. | Episodes/frequency of caregiver engagement | How many interactions did caregivers have with the platforms? How frequent were the interactions? |
| Aim of study | Describe aim as per author's description. | Tools used for data collection | Which digital tools were used to collect data? |
| Participant ages | What age(s) were the participating children? | Duration of intervention | How long did the intervention last? |
| Involvement of caregivers or guardians | Were caregivers involved? If so did they: 1) give consent 2) submit data 3) assist the child only 4) other, if so, describe. | Blinding | Was there any blinding? |
| SES/Race/Gender of participants | List, where provided. | Pushes/reminders | Were reminders used to retain participants? If so, describe. How many and how often? |
| Steering/advisory group formation | Was a steering group or advisory group formed to guide the study? If so, describe. | Attrition rates | What were the attrition rates of participants? |
| Process of randomisation | How were participants randomised? List the tools and methods used. | Outcomes relevant to the review | To be decided. |
| Context | Were participants of a specific group recruited? If so, list. | Limitations | What study limitations do the authors cite? |
| Platforms used for recruitment and retention | List all digital platforms used for recruitment and retention. | Funding | Where did the funding for the project come from? |

Description column is for reviewer clarification during data charting.

### Collating, Summarising, and Reporting Results

We will conduct a risk of bias (RoB) assessment of included studies using the RoB1 tool outlined in the Cochrane Handbook for Systematic Reviews of Interventions [31]. This tool assesses selection bias, performance bias, detection bias, attrition bias, reporting bias, and other biases. To decrease the time spent assessing RoB, we will use RobotReviewer from Vortext Systems [32]. This freely available software, uses machine learning to semi-automate RoB assessments. RobotReviewer has shown an ability to decrease the workload of large reviews [33]. The creators of RobotReviewer stipulate that although machine learning can be extremely useful, it should be validated by the people using it [33]. We will validate results with two reviewers manually assessing RoB from the five original studies used for the extraction pilot and compare them to those from RobotReviewer [32]. Once we reach an IRR of 0.75 using Cohen’s kappa [30], a single reviewer will complete the remaining studies’ RoB.

We will summarise the charted data and present it in the text of the full report and in a tabular fashion as it addresses the review’s objectives. Representations of digital tools and platforms used for each phase of the trials, participants’ ages and characteristics, follow-up or retention methods used, consent verification methods used, and results of interest will likely be best presented in graphical form. However, because of the iterative nature of this review and breadth of the studies, other forms of data depiction may develop in the final report. It is notable that the aim of a scoping review is not to report or analyse the results of included studies and that we will collate results as they apply in the context of the review question.

## Discussion

This scoping review aims to identify and characterize how online randomised trials with children have been designed and implemented, and identify the gaps in the research. The review will employ a thorough and robust search strategy, chart the desired data, and summarise it in order to establish how these trials are managed. Our aim is that this review will allow for recommendations for future research directions and help guide researchers to improve the execution of these trials.

The challenges faced by researchers in these types of trials differ depending on the participants’ ages, the interventions used, and the primary objectives of the studies. Sensitivity, technological knowledge, and forethought are required to protect the child and find the appropriate digital tools for recruitment. In trials where maintaining the child’s autonomy is critical, it can be a complex balance also to involve the caregivers to ensure legal and ethical integrity. Collecting and validating unique, individual data sets digitally requires additional verification steps compared to what a centralised trial might demand. Finally, and arguably most importantly, confirming informed consent and assent from both caregivers and children online calls for transparency, caution, and creativity in some cases. These additional challenges can be daunting for researchers to tackle; however, trialists have made innovative advances in online research with children in the last decade.

Our interest in this field stems from the ability of decentralised trials to reach a larger participant base, potentially decrease research waste, and expand inclusivity in paediatric trials. However, we acknowledge that internet-based trials inherently exclude some populations. There remains a digital divide through either disadvantage, infrastructure availability, or personal choice. We have chosen to focus on entirely online, decentralised trials to describe their benefits and challenges, and it is outside the scope of this work to address and include trials that are not internet-based.

## Supporting information

S1 Checklist

## Data Availability

All search result files are available from Open Science Forum (https://osf.io/gfp42/). All other relevant data are within the manuscript and its Supporting Information Files.

https://osf.io/ha3kf

## Acknowledgements

This work contributes to a doctoral project for one of the authors (SL).

## Supporting Information

**S1 Checklist. Preferred Reporting Items for Systematic Reviews and Meta-Analyses Extension for Scoping Reviews (PRISMA-ScR) Checklist**. Grey-filled rows do not apply during the protocol stage.

**S2 Appendix. Search strategy developed in MEDLINE (Ovid) for “Online randomised trials with children: A scoping review protocol”**. Search conducted February 16 2022.

