## Supplementary material for "Online randomised trials with children: A scoping review protocol": S1 Checklist

### S1 Checklist. Preferred Reporting Items for Systematic Reviews and Meta-Analyses Extension for Scoping Reviews (PRISMA-ScR) Checklist

| **SECTION** | **ITEM** | **PRISMA-ScR CHECKLIST ITEM** | **REPORTED ON PAGE #** |
| --- | --- | --- | --- |
| **TITLE** | | | |
| Title | 1 | Online Randomised Trials with Children: A Scoping Review Protocol | 1 |
| **ABSTRACT** | | | |
| Structured summary | 2 | Abstract provided | 2 |
| **INTRODUCTION** | | | |
| Rationale | 3 | This scoping review will identify, describe, and characterize how online, decentralized trials have been conducted with children in order to understand how they may be most effectively employed. | 2, 3 |
| Objectives | 4 | The objectives of this scoping review are: (a) to determine what methods and tools have been used to create and conduct online trials with children and (b) to identify the gaps in the knowledge in this field.  The population is children. The context is randomised and quasi-randomised trials. The concept is internet-based interventions. | 3 |
| **METHODS** | | | |
| Protocol and registration | 5 | This protocol statement and registration at Open Science Forum. | <https://osf.io/ha3kf> |
| Eligibility criteria | 6 | All randomised and quasi-randomised trials with children published in English conducted entirely online. | 5 |
| Information sources* | 7 | Databases: MEDLINE, CENTRAL, CINAHL, Embase  Registries: ICTRP, EU Clinical Trials Register, NIH Clinical Trials Register  Preprints: medRxiv, JMIR Preprints, HRB Open Research, and Advance from SAGE  Internet Searches with expert consultation | 4 |
| Search | 8 | Search strategy provided for MEDLINE (Ovid) | S2 Appendix |
| Selection of sources of evidence† | 9 | Pilot tests with two independent reviewers will be conducted in order to achieve an inter-rater reliability of 0.75 using Cohen’s kappa. A single reviewer will perform remaining screenings. | 4, 5 |
| Data charting process‡ | 10 | Preliminary data charting form provided and to be piloted and revised by two independent reviewers. | Table 1 |
| Data items | 11 | 1. Author 2. Title 3. Year of Publication 4. Country(ies) of Conduct 5. Country(ies) of Participants 6. Type of study 7. Type of publication 8. Aim of study 9. Participant ages 10. Involvement of caregivers or guardians 11. SES/Race/Gender of participants 12. Steering/advisory group formation 13. Process of randomisation 14. Context 15. Platforms used for recruitment and retention 16. Cross-platform compatibility 17. Tools used for data protection processes 18. Tools used for consent and assent 19. Consent/assent validation 20. Interventions 21. Episodes/frequency of engagement 22. Episodes/frequency of caregiver engagement 23. Tools used for data collection 24. Duration of intervention 25. Blinding 26. Pushes/reminders 27. Attrition rates 28. Outcomes relevant to the review 29. Limitations 30. Funding | Table 1, pp 6, 7 |
| Critical appraisal of individual sources of evidence§ | 12 | We will be assessing Risk of Bias (RoB) using the RoB1 tool from the Cochrane Handbook. To assist in the RoB analysis we will use RobotReviewer software from Vortext Systems. RoB will be independently assessed by two reviewers for five studies, once a Cohen’s kappa inter-rater reliability of 0.75 is reached, a single reviewer will complete the RoB analysis. | 7 |
| Synthesis of results | 13 | We will synthesize the data in text, tabular and graphical formats. Other forms of data depiction may evolve iteratively during the data collating process. | 7 |
| **RESULTS** | | | |
| Selection of sources of evidence | 14 |  |  |
| Characteristics of sources of evidence | 15 |  |  |
| Critical appraisal within sources of evidence | 16 |  |  |
| Results of individual sources of evidence | 17 |  |  |
| Synthesis of results | 18 |  |  |
| Summary of evidence | 19 | The data will be summarized in the context of the review objectives, focusing on the digital technologies leveraged in randomised, online trials with children. The focus will be on what technologies and methods were beneficial or challenging, where the gaps in the research are, and inform how to manage these trials successfully in the future. | 7 |
| Limitations | 20 | This scoping review will only include and assess decentralized, online randomised trials. There are many types of decentralized trials that are ‘hybird’, which are effective and will not be included. | 7 |
| Conclusions | 21 |  |  |
| **FUNDING** | | | |
| Funding | 22 | This research was supported by the Health Research Board [TMRN-2021-001] and by the College of Medicine, Nursing and Health Sciences at NUI Galway. | 8 |

JBI = Joanna Briggs Institute; PRISMA-ScR = Preferred Reporting Items for Systematic reviews and Meta-Analyses extension for Scoping Reviews.

* Where *sources of evidence* (see second footnote) are compiled from, such as bibliographic databases, social media platforms, and Web sites.

† A more inclusive/heterogeneous term used to account for the different types of evidence or data sources (e.g., quantitative and/or qualitative research, expert opinion, and policy documents) that may be eligible in a scoping review as opposed to only studies. This is not to be confused with *information sources* (see first footnote).

‡ The frameworks by Arksey and O’Malley (6) and Levac and colleagues (7) and the JBI guidance (4, 5) refer to the process of data extraction in a scoping review as data charting*.*

§ The process of systematically examining research evidence to assess its validity, results, and relevance before using it to inform a decision. This term is used for items 12 and 19 instead of "risk of bias" (which is more applicable to systematic reviews of interventions) to include and acknowledge the various sources of evidence that may be used in a scoping review (e.g., quantitative and/or qualitative research, expert opinion, and policy document).

*From:* Tricco AC, Lillie E, Zarin W, O'Brien KK, Colquhoun H, Levac D, et al. PRISMA Extension for Scoping Reviews (PRISMAScR): Checklist and Explanation. Ann Intern Med. 2018;169:467–473. [doi: 10.7326/M18-0850](http://annals.org/aim/fullarticle/2700389/prisma-extension-scoping-reviews-prisma-scr-checklist-explanation).
