## Supplementary material for "Online randomised trials with children: A scoping review protocol": S2 Appendix

### **S2 Appendix. Search strategy developed in MEDLINE (Ovid) for “Online randomised trials with children: A scoping review protocol” ( Conducted 16 February 2022).**

Ovid MEDLINE(R) ALL <1946 to February 16, 2022>

| 1 | (toddler$ or preschool$ or pre-school$ or preschool child$ or kindergarten$ or kinder-garten$ or child$ or kid$ or boy$ or girl$ or pediatric$ or paediatric$ or schoolage$ or school-age or school age$ or pre-teen$ or adolescen$ or teen$ or youth$ or young m#n$ or young wom#n$).tw,kw. |
| --- | --- |
| 2 | Child, Preschool/ |
| 3 | exp Child/ |
| 4 | Adolescent/ |
| 5 | or/1-4 |
| 6 | ((Internet$ or online$ or on-line$ or web$ or computer$ or digital$ or virtual$ or remote$ or decentrali$) adj3 (deliver$ or trial$ or intervention$ or study or studies or random$ or rct$)).tw,kw. |
| 7 | ((internet$ or online$ or on-line$ or web$ or computer$ or laptop or ipad or i-pad or digital$ or virtual$ or remote$ or technolog$ or software or smartphone$ or smart-phone$ or smart phone$ or mobile or iPhone or Android or cellphone$ or cell-phone$ or cell phone$ or social media or twitter or facebook or Instagram or snapchat) adj3 (application$ or app$ or deliver$ or intervention$ or treat$ or therap$ or program$)).tw,kw. |
| 8 | (ehealth or e-health or electronic health or mhealth or m-health or mobile health or telehealth or tele-health or telemedicine or tele-medicine).tw,kw. |
| 9 | Internet-Based Intervention/ |
| 10 | Therapy, Computer-Assisted/ |
| 11 | or/7-10 |
| 12 | 6 and 11 |
| 13 | randomized controlled trial.pt. |
| 14 | controlled clinical trial.pt. |
| 15 | randomi#ed.ab. |
| 16 | placebo$.ab. |
| 17 | drug therapy.fs. |
| 18 | randomly.ab. |
| 19 | trial.ab. |
| 20 | groups.ab. |
| 21 | or/13-20 |
| 22 | exp animals/ not humans.sh. |
| 23 | 21 not 22 |
| 24 | 5 and 12 and 23 |
